# Challenging deficient inhibitory conditioned pain modulation as common chronic pain feature and detectable subgroup characteristic

**DOI:** 10.64898/2026.05.01.26352197

**Authors:** Laura Sirucek, Iara De Schoenmacker, Lindsay Gorrell, Robin Lütolf, Anke Langenfeld, Florian Brunner, Jan Rosner, Mirjam Baechler, Brigitte Wirth, Michèle Hubli, Petra Schweinhardt

## Abstract

Deficient descending pain inhibition assessed by conditioned pain modulation (CPM) is considered a common feature of various chronic pain disorders. Typically, CPM studies focus on one particular disorder making direct comparisons between disorders difficult. This cross-sectional study aimed to compare CPM effects between three clearly distinct chronic pain disorders and pain-free controls. Furthermore, patients were pooled with controls to explore whether subgroups showing different CPM effects could be separated independent of cohort membership. One hundred and forty participants (patients: 53 non-specific chronic low back pain [nsCLBP], 15 complex regional pain syndrome [CRPS], 14 neuropathic pain after spinal cord injury [painSCI]; 58 controls) were included. CPM effects were assessed in a remote, pain-free area using pressure pain thresholds as test stimulus and a cold water bath as conditioning stimulus. Cohort differences in CPM effects were analyzed using linear mixed models. The presence of subgroups showing different CPM effects was tested using latent class linear mixed models. CPM effects differed between cohorts (p = 0.011), driven mainly by reduced inhibitory CPM effects in patients with nsCLBP compared to patients with painSCI. Latent class analysis detected 3 subgroups with varying degrees of significant inhibitory CPM effects (p’s ≤ 0.002). All subgroups comprised patients and controls. These results oppose deficient descending pain inhibition as a common feature of chronic pain disorders. Additionally, the failure to identify subgroups without inhibitory CPM effects within a heterogenous patient/control sample challenges the utility of deficient CPM as predictor of chronic pain or treatment efficacy.

**Perspective:** Inhibitory conditioned pain modulation, a measure of descending pain inhibition, is not consistently impaired across distinct chronic pain disorders. Furthermore, identifying individuals with impaired conditioned pain modulation within a heterogenous sample is difficult. Thus, for conditioned pain modulation to be clinically useful, its variability needs to be better understood.

## 1. Introduction

Over the past decades, chronic pain prevalence has continuously increased [1,72,88], emphasizing the growing need for advancements in chronic pain management. Currently available treatment strategies show limited success [33] which is partly due to an insufficient understanding of chronic pain mechanisms. If, for a given patient, the predominant chronic pain mechanism was known, mechanism-based instead of one-size-fits-all treatment approaches could be pursued.

It has been proposed that pain modulatory profiles could inform treatment approaches [23,84]. One of the suggested key pain modulatory measures is conditioned pain modulation (CPM) [82]. CPM is considered the human analogue of diffuse noxious inhibitory controls [42,43] and serves as a measure of descending pain modulatory capacities. In CPM paradigms, the modulatory effect of a noxious conditioning stimulus (CS) on another, remotely applied, noxious test stimulus (TS) is assessed [82]. Typically, the CS inhibits the pain perceived in response to the test stimulus [48,69]. A reduction of such pain-inhibitory CPM effects (herein referred to as deficient CPM) has been reported in various chronic pain disorders [44,84], indicating a dysfunctional descending pain inhibitory system. However, a direct comparison between chronic pain disorders is difficult because CPM studies typically focus on only one particular disorders at a time [44] and use different CPM paradigms associated with different CPM effects [31,37,52,53,56].

Therefore, the first aim of this study was to compare CPM capacities measured by the same CPM paradigm between patients with clearly distinct chronic pain disorders recruited from our multidisciplinary clinical environment and pain-free controls. A standard CPM paradigm [53,61] with pressure pain thresholds (PPTs) as TS and a cold water bath as CS was applied in a pain-free body area of patients with non-specific chronic low back pain (nsCLBP), complex regional pain syndrome (CRPS) and neuropathic pain after spinal cord injury (painSCI), and pain-free controls. It was hypothesized that all chronic pain cohorts would present with reduced inhibitory CPM effects compared to pain-free controls. Additionally, associations of CPM effects with clinical pain characteristics and psychological factors were explored because previous studies have shown that higher clinical pain intensity [47], longer pain duration [17,47], larger spatial pain extent [18,26], as well as certain psychological factors [4,51] are associated with deficient CPM. It was expected that more severely impaired patients would present with reduced inhibitory CPM effects.

Deficient CPM has furthermore been suggested to be a predictive factor for chronic pain development [10,83] and analgesic efficacy of pharmacological interventions [11,85]. Yet, for CPM effects to serve as such a predictor in a clinical setting, it has to be possible to identify individuals with deficient CPM within a larger, heterogenous population. Thus, the second aim of this study was to explore whether within the diverse sample of patients with distinct chronic pain disorders and pain-free controls, subgroups showing different CPM effects could be identified. For that, latent class linear mixed models were performed on the full dataset, to identify potential homogenous subgroups independent of cohort membership.

## 2. Methods

Please note that substantial material in this section is identical to a manuscript focusing on the CPM data of the patients with nsCLBP and the respective controls in association with magnetic resonance spectroscopy of the periaqueductal gray [73]. The CPM data of the patients with painSCI included in the present study have been previously published elsewhere [46].

### 2.1. Participants

One hundred four patients with chronic pain (64 patients with nsCLBP, 21 patients with CRPS, 19 patients with painSCI) and 64 individually age- and gender-matched, pain-free controls (between 18 and 80 years of age) were consecutively recruited between November 2019 and April 2022 (with a COVID-19 pandemic-related recruitment break from March to May 2020). Patient recruitment was performed at the Balgrist University Hospital and for patients with nsCLBP, additionally via advertisements in Swiss Chiropractic practices and patient magazines. Controls were recruited via online advertisements and oral communication. Given that, to the best of our knowledge, this is the first study comparing CPM effects between three clearly distinct chronic pain disorders and pain-free controls, sample sizes were chosen based on previous disorder-specific CPM studies that included both patients and controls [2,26,39,47]. Data were collected between November 2019 and April 2022. Gender information was gathered via self-report allowing for the choice between “male”, “female”, and “diverse”. Study participation was open to all individuals meeting the inclusion and exclusion criteria independent of race, ethnicity, or gender. Specific inclusion criteria for the chronic pain cohorts were:

- nsCLBP: LBP as the primary pain complaint of a duration longer than 3 months without signs of serious underlying pathology (e.g., signs of infection, fractures, inflammation) or signs of symptomatic radiculopathy (motor and sensory deficits).
- CRPS: Fulfillment of the clinical Budapest criteria [28] at inclusion as diagnosed by an experienced rheumatologist (FB) and pain of a duration longer than 3 months.
- painSCI: Thoracolumbar (below C8) SCI of a duration longer than 1 year and the presence of neuropathic pain according to current diagnostic criteria [14].

Exclusion criteria for all patient cohorts comprised any major medical, neurological or psychiatric disorder other than nsCLBP/CRPS/painSCI, symptomatic radiculopathy (i.e., motor and sensory deficits), pregnancy, or inability to follow study instructions. For controls, the same exclusion criteria were applied with the addition that controls were required to present without history of chronic pain and be free of low back pain lasting longer than 3 consecutive days during the last year. The study was approved by the local ethics committee “Kantonale Ethikkommission Zürich” (Nr.: PB_2019-00136, PB_2016-02051 and EK-04/2006), registered on clinicaltrials.gov (NCT04433299 and NCT02138344), and performed according to the guidelines of the Declaration of Helsinki (2013). Written informed consent was obtained from all participants before the start of the experiment.

Patients and members of the public were not involved in the present study’s conceptualization, design, conduct, or dissemination of the study results. We intend to disseminate the main results to all participants.

### 2.2. Study design

This study was part of a larger project (Clinical Research Priority Program “Pain”, https://www.crpp-pain.uzh.ch/en.html) which was comprised of 3 experimental sessions of approximately 3 hours each and electronic questionnaires. In the first 2 sessions, participants underwent clinical, neurophysiological and psychophysical assessments. The CPM assessment was always performed in the second session preceded by the acquisition of pain-related evoked potentials. The third session comprised the acquisition of magnetic resonance data. The present study concerns the CPM data, pain drawings (and for patients with nsCLBP, pain provocation patterns) acquired during the clinical assessment, and questionnaire data.

### 2.3. Conditioned pain modulation

Participants were tested in a prone (nsCLBP), supine, or seated (CRPS and painSCI; based on the patient’s preference) position in a quiet room (temperature 20-25 °C). The CPM assessment was performed by a trained experimenter. Participants were given standardized verbal instructions.

The TS of the CPM was applied at the patient’s most painful area and at a remote, pain-free control area. The order of the 2 areas was randomized and counterbalanced. In the present study, only data from the remote area were used as the patients’ most painful areas were highly heterogenous and differed in terms of sensibility, e.g., in patients with painSCI, which would hamper comparability between cohorts. The remote area was in most cases the hand, specifically the non-dominant hand for patients with nsCLBP and the hand contralateral to the most painful area for patients with CRPS and painSCI. Exceptions were patients with CRPS affecting the hand, because it has been shown that symptoms can spread to the contralateral limb [64]. In these patients, the shoulder contralateral to the most painful area served as the remote area. The tested remote area in each control was identical to the patient to whom they were individually age- and gender-matched.

The CS consisted of a circulating cold water bath (9 ± 0.5 °C) in which the participants immersed the hand contralateral to the remote area up to the wrist for 2 min. Patients with CRPS affecting the hand immersed their ipsilateral foot up to the ankle. The perceived pain intensity of the CS was rated on a numeric rating scale (NRS) from 0 ‘No pain’ to 10 ‘Maximum pain tolerable’ directly after immersion, 30 s after immersion and immediately before withdrawal of the hand or foot.

The TS consisted of PPTs assessed before, during (30 s after CS onset; ‘parallel CPM effect’), and after (’sequential CPM effect’) the CS. PPTs were chosen as TS because (1) deep afferents are suggested to be subject to stronger descending pain inhibition compared to superficial afferents [86,87] and (2) its combination with a cold water bath as CS has been shown to be the CPM paradigm with the highest intra-session reliability [53] and with larger inhibitory CPM effects compared to other TS-CS combinations [31]. Pressure was applied using a hand-held mechanical algometer (Wagner Instruments, Greenwich, CT, USA) with a circular rubber tip (1 cm diameter). PPTs were assessed at the thenar eminence (if remote area = hand) or over the deltoid muscle (if remote area = shoulder). One PPT per timepoint was determined using the method of limits [67]. If no pain was reported at 10 kg/cm^2^ (safety cut-off to avoid tissue damage), a value of 11 kg/cm^2^ was assigned as the PPT. Besides PPTs, the CPM paradigm comprised three additional TS’s, i.e., pressure temporal summation of pain (TSP), heat pain thresholds and heat TSP. The order of pressure and heat assessments was randomized and TSP was always performed after the pain threshold assessments. These TS’s will not be discussed; relevant here is that the paradigm was identical between the patients and the matched controls and that the TS order was randomized.

To test whether the used CPM paradigm induced a ‘true’ CPM effect beyond influences unrelated to the painfulness of the CS (e.g., repeated-measures effects) [37], a CPM_SHAM_ paradigm was implemented. The CPM_SHAM_ paradigm was performed identically to the CPM paradigm except for an ambient temperature water bath (32 ± 0.5 °C) as CS. The order of the CPM and CPM_SHAM_ paradigms was randomized with a minimum of 10 min between. Data reporting on the CPM_SHAM_ paradigm was not included to answer the present research questions, because adding an extra independent variable (i.e., type of paradigm) to the linear mixed models was not suitable given the sample size. Nevertheless, data from the CPM_SHAM_ paradigm were used to approximate the standard error of measurement (SEM) for PPT and to classify patients into CPM-inhibitors, CPM-facilitators and CPM-non-responders (see Section 2.6.3).

### 2.4. Clinical pain characteristics and psychological factors

As part of the electronic questionnaires, patients reported their average clinical pain intensity over the past 4 weeks (NRS: 0 ‘No pain’ to 10 ‘Maximum pain’) which was considered the patients’ ‘trait’ pain. In addition, patients reported the current pain intensity in their most painful area before the first water bath of the CPM paradigm on the same NRS used for the cold water bath pain (0 ‘No pain’ to 10 ‘Maximum pain tolerable’). This *within-CPM-session clinical pain* was considered the patients’ ‘state’ pain.

Patients completed pain drawings before the first experimental session to assess the spatial pain extent of their typically painful body areas [9,68]. Participants were instructed to shade their typically painful areas on printed standardized body charts (frontal and dorsal view). After manual contouring of the shaded areas, the pain drawings were digitalized and processed using a custom-made software that calculated the spatial extent of the shaded areas as a percentage of total body area. Only disorder-associated areas (i.e., between the twelfth rib and the gluteal fold and in the legs for patients with nsCLBP, syndrome-specific areas for patients with CRPS, and neuropathic pain areas for patients with painSCI) were considered for further analyses.

Participants also completed the Pain Catastrophizing Scale (PCS) [75] and the Hospital Anxiety and Depression Scale (HADS) [89]. The PCS consists of 13 items with a score range between 0 and 52 (> 30 clinically relevant level of catastrophizing). The HADS consists of 14 items with a 7-item anxiety-subscale and a 7-item depression-subscale, with a score range between 0 and 21 per subscale (8-10 moderate and 11-21 high probability for a mood disorder).

For the nsCLBP cohort, the contribution of nociceptive, neuropathic and nociplastic mechanisms [71] to the patients’ pain was estimated. For that, information on pain provocation patterns (tested during the clinical assessment based on an evidence-based diagnostic classification system for low back pain [77,78]), scores from neuropathic pain questionnaires (i.e., painDETECT [16]), and pain widespreadedness was used. Clear discogenic/facetogenic/sacroiliac provocation patterns, painDETECT scores ≤ 12, and a localized pain extent were considered indicators of nociceptive pain mechanisms; positive slump test, reduced patella or achilles tendon reflexes, hyposensitivity to light touch), muscle weakness, painDETECT scores ≥ 19, and neuroanatomically plausible dermatomal pain extent were considered indicators of neuropathic pain mechanisms; and inconsistent pain provocation patterns and widespread pain ( i.e., axial pain in combination with upper and lower limb pain, as well as left and right limb pains, according to [29,81] and based on painful areas indicated in the Widespread Pain Index [81]) were considered indicators of nociplastic pain mechanisms.

### 2.5. Confounding factors

Prior to the CPM session, information about the menstrual cycle phase was obtained if applicable, i.e., in premenopausal women not using menstruation-suppressing contraceptives. In the electronic questionnaires, participants were asked to indicate any regular medication intake. Medications were classified according to the ATC/DDD classification by the World Health Organization (http://www.whocc.no/atc_ddd_index/). The following categories were considered pain-relevant: M01A (anti-inflammatory and anti-rheumatic drugs and non-steroids), N02 (analgesics), N03 (antiepileptics), N05 (psycholeptics), and N06 (psychoanaleptics). Information on pain-relevant medication intake was dichotomized (yes/no) for further statistical analyses.

### 2.6. Statistical analysis

All statistical analyses were performed in RStudio [76] for Mac (2022.12.0+353). Statistical significance was set at *α* = 0.05. All post-hoc tests were false discovery rate (FDR)-corrected. For exploratory analyses, i.e., associations of CPM effects with clinical pain characteristics and psychological factors, and analyses of potential confounding factors, no multiple comparison correction was applied because false negatives were considered less desirable than false positives. Depending on the statistical test used, raw values or model residuals were assessed for normal distribution using inspection of histograms and QQplots. All ordinal or non-normally distributed variables are reported as median (interquartile range) and were analyzed using non-parametric tests. All continuous and normally distributed variables are reported as mean (standard deviation) and were analyzed using parametric tests. Missing data was excluded from analyses except for linear mixed models, which can account for missing data.

CPM effects were calculated as follows: parallel CPM effect = PPT during – PPT before, sequential CPM effect = PPT after – PPT before. Thus, inhibitory CPM effects (i.e., PPT increases) are denoted by a positive value and facilitatory CPM effects (i.e., PPT decreases) by a negative value.

CPM effects were analyzed using linear mixed models with 2 different approaches. The first approach (cohort-specific) addressed the present study’s first aim, i.e., to compare CPM effects between chronic pain disorders and pain-free controls. The second approach (subgroup-specific) addressed the present study’s second aim, i.e., to explore whether within the full dataset, homogenous subgroups showing different CPM effects can be identified. The 2 approaches are described in detail in the sections below.

For both approaches, the following applied:

- Participant identifier was included as random intercept effect.
- Potential influences of age and gender on the model’s dependent variable were examined by testing for age and gender main effects and interaction effects with the model’s independent variables. The models were always fitted with all interactions first, except for ‘age X gender’ interactions given the low number of participants of different ages within a certain gender. If the age/gender interactions were not significant, they were removed from the model to assess age/gender main effects. If the age/gender main effects were not significant, they were also removed from the model. Of note, the effects of interest (indicated in the model descriptions below) were always left in the model when assessing age/gender influences.

If assumptions of linear mixed models were not met, a Friedman test with post-hoc Wilcoxon signed rank tests was used to analyze CPM effects. The influence of age and gender was assessed separately with Spearman correlations and Wilcoxon rank sum tests with PPT changes (i.e., during - before and after - before) as dependent variable to assess age/gender interaction effects, and if not significant, the average of PPTs before, during, and after CPM, as dependent variable to assess age/gender main effects.

The following effect sizes were calculated for the respective statistical tests:

- Linear models: partial η^2^ (small: 0.01, medium: 0.06, large: 0.14) [5]
- Kruskal-Wallis tests: η^2^ (using benchmarks for partial η^2^ although these have not been officially established for η^2^: 0.01, medium: 0.06, large: 0.14) [65]
- Post-hoc t-tests: Cohen’s d (small: < 0.5, medium: 0.5-0.8, large: > 0.8) [5]
- Post-hoc Wilcoxon rank-sum tests: r (small: 0.1-< 0.3, medium: 0.3-< 0.5, large: ≥ 0.5) [6]

No effect sizes are reported for linear mixed models because no agreement on standard effect sizes exists [66].

#### 2.6.1. Cohort-specific approach

The presence of cohort differences in CPM effects was tested using a linear mixed model (R package ‘nlme’, function ‘lmer’) with PPTs as the dependent variable and the independent variables timepoint (3 levels: ‘before’, ‘during’, ‘after’), cohort (4 levels: ‘nsCLBP’, ‘CRPS’, ‘painSCI’, ‘controls’) and the interaction of interest, i.e., ‘timepoint X cohort’ (PPT model). Such a model is highly dependent on PPTs before the CS (herein referred to as baseline PPTs). In an attempt to reduce the influence of baseline PPTs, the same analysis was performed with the PPT *changes* (i.e., during - before and after - before) as the dependent variable and baseline PPT as additional independent variable (PPT *change* model; for details see Supplementary Methods M1).

Differences in baseline PPTs, i.e., baseline pressure pain sensitivity, between the cohorts were tested using a Kruskal-Wallis test and post-hoc pairwise Wilcoxon rank-sum tests with continuity correction.

To examine whether each cohort showed significant CPM effects, four linear mixed models (or Friedman tests if assumptions for linear mixed models were not met) were performed for each cohort separately with PPTs as the dependent variable and timepoint as the independent variable. Post-hoc tests were performed using planned comparisons (timepoint during vs. timepoint before, timepoint after vs. timepoint before).

All cohort-specific models were assessed for model residuals, i.e., single datapoints, which exerted large influence on the model, so-called influential cases [13]. Influential cases were identified based on studentized residuals (R function rstudent()) and Cook’s distance [13]. For studentized residuals, a cut-off of 1.96 was chosen [13]. For Cook’s distance, a conservative cut-off of 4/(n-k-1) [27] (with n = number of observations and k = number of predictors in the model) was chosen. If influential cases were identified, the models were re-run without the influential cases to examine whether the statistical inference of the model changed. This ensured that obtained results were not driven by a small number of extreme datapoints. If not otherwise stated, removal of influential cases did not change the statistical inferences and the results of the full data set are reported.

##### Clinical pain characteristics, psychological factors and associations with CPM effects

Kruskal-Wallis tests were used to compare clinical pain characteristics between chronic pain disorders. Investigated clinical pain characteristics were: (i) average clinical pain intensity over the past 4 weeks, (ii) pain duration (in months), (iii) spatial pain extent, and (iv) within-CPM-session clinical pain because ‘state’ pain can confound comparisons between pain and pain-free cohorts. [3] Post-hoc tests were performed using pairwise Wilcoxon rank-sum tests with continuity correction. The same approach was followed to compare PCS and HADS anxiety and depression scores between cohorts (including pain-free controls).

Associations of CPM effects (parallel and sequential) with the 4 clinical pain characteristics and 3 psychological factors were examined using Spearman correlations. This was performed in each chronic pain disorder independently and for all chronic pain disorders combined. After that, rank-based regressions [35] were used to assess whether the observed correlations differed between the cohorts. In these regressions (N=14), the dependent variable was the CPM effect (parallel or sequential) and the independent variables were the respective clinical pain characteristic or psychological factors, and cohort. The effect of interest was the ‘pain characteristic/psychological factor X cohort’ interaction. Given that rank-based regressions are robust against outliers in the response space [35], no assessment of influential cases was performed.

#### 2.6.2. Subgroup-specific approach

The presence of subgroups showing different CPM effects was tested using latent class linear mixed models [60] (R package ‘lcmm’, function ‘hlme’). These models enable the detection of subgroups showing discrete trajectories in a variable over time [45,58], in this case, PPT changes throughout the CPM paradigm. As above, 2 models were investigated, 1 with PPTs as dependent variable (PPT model) and 1 with PPT *changes* as dependent variable (PPT *change* model). The models were set up as above, except that the cohort was not included as an independent variable. Age and gender influences were tested in the single-class model which served as the basis for the multi-class models. Latent class membership was assessed based on differences in slopes between timepoints (mixture = ∼ timepoint). The estimation algorithm was initialized with values resulting in the best log-likelihood after a specified (here 15) number of iterations from a number of random sets (here 100) of initial values. The number of possible classes, i.e., subgroups, was increased until the lowest Bayesian Information Criterion (BIC) [63,70] was achieved and a further increase in the number of subgroups did not further improve the BIC. Additionally, the model needed to present high confidence of subgroup membership (> 80%) and a minimum subgroup size of 5% of the sample [58,63].

To examine whether each subgroup identified in the PPT model (PPT-subgroup) showed significant CPM effects, a linear mixed model (or a Friedman test if assumptions for linear mixed models were not met) was performed for each PPT-subgroup separately with PPTs as the dependent variable and timepoint as the independent variable. Post-hoc tests were performed using planned comparisons (’timepoint during’ vs. ‘timepoint before’, ‘timepoint after’ vs. ‘timepoint before’).

Additionally, to analyze whether PPT-subgroups indeed differed in their CPM effects and whether a difference was present in parallel or sequential CPM effects, 2 additional linear models were performed with PPT change (parallel or sequential) as the dependent variable and subgroup membership as the independent variable. Post-hoc tests were performed using pairwise t-tests.

#### 2.6.3. CPM responder classification

In addition to the cohort-specific and subgroup-specific approaches, all participants were classified as CPM-inhibitors, CPM-facilitators and CPM-non-responders based on the SEM of PPT [34]. The SEM was calculated using CPM_SHAM_ data of all included controls which were randomized to perform the CPM_SHAM_ paradigm first (N = 36) with the formula [34]:

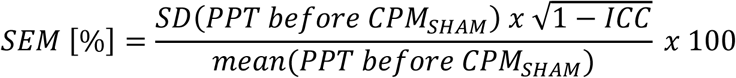

The intraclass correlation coefficient (two-way, absolute agreement, single rater/measurement) was calculated between the PPT before and the PPT during CPM_SHAM_. Participants were classified as CPM-inhibitors or CPM-facilitators if in the CPM paradigm, they presented with a relative PPT increase or PPT decrease > 2 SEM, respectively, and otherwise as CPM-non-responders [34]. Relative PPT increases or decreases were calculated as (absolute PPT increase or decrease / PPT before)*100. The maximum possible PPT increase or decrease was set to ±100%. Proportions of CPM-inhibitors, CPM-facilitators and CPM-non-responders were compared between chronic pain cohorts and controls, as well as between the identified subgroups, using Fisher’s exact tests.

#### 2.6.4. Confounding factors

Proportions of the different menstrual cycle phases (menstruation, follicular, ovulation, or luteal) and of participants with and without regular pain-relevant medication intake were compared between chronic pain cohorts and controls using Fisher’s exact tests. If proportions were similar across cohorts, data were pooled for analyses on whether menstrual cycle phase/regular pain-relevant medication intake had an influence on CPM effects using Kruskal-Wallis tests.

## 3. Results

### 3.1. Participants

Of the 168 recruited individuals, 11 were excluded (3 (nsCLBP) cancelled prior to the first session, 3 (2 painSCI, 1 nsCLBP) showed signs of neurological disorders (other than SCI), 1 (nsCLBP) showed indications of a psychiatric disorder, 2 (controls) showed abnormal sensory findings, 1 (CRPS) discontinued testing due to discomfort and 1 (control) developed low back pain between the time of inclusion and testing). Of the remaining 157 participants, 2 (CRPS) had pain in both hands and shoulders, i.e., lacked a pain-free remote area, 1 (CRPS) did not complete CPM testing, 1 (CRPS) presented with CRPS type 2 and was excluded to increase homogeneity within the cohort, and 13 (6 nsCLBP, 1 CRPS, 3 painSCI and 3 controls) did not tolerate the full 2 min cold water bath (which could hamper comparability) and were also excluded.

Thus, the final sample comprised 140 participants (all Europeans), including 53 patients with nsCLBP, 15 patients with CRPS, 14 patients with painSCI and 58 controls. The participant demographics, clinical pain characteristics, and psychological factors are described in Table 1.

**Table 1.**
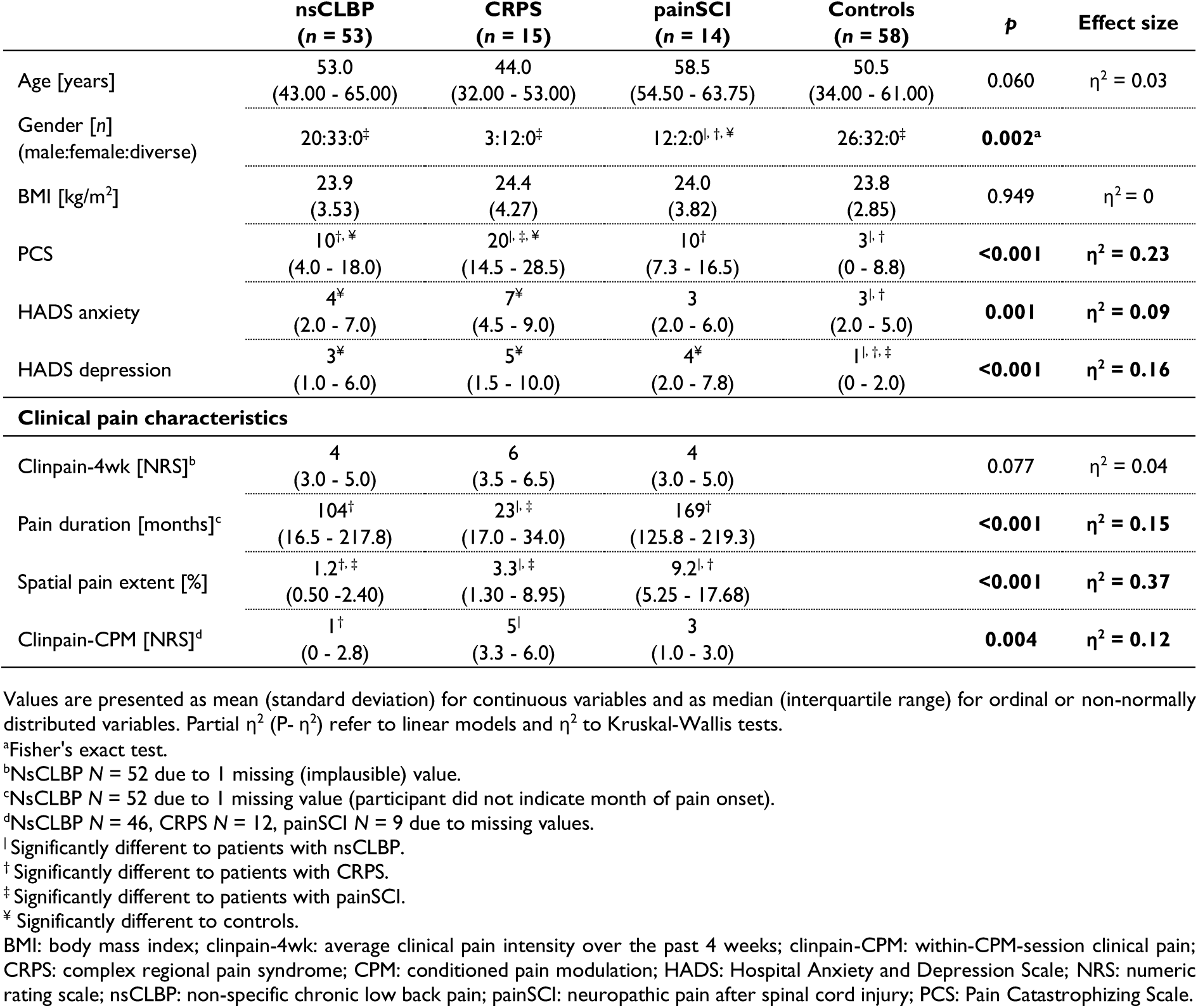
Participant demographics and clinical characteristics.

Of the patients with nsCLBP, 33/53 (62.3%) showed clear discogenic/facetogenic/sacroiliac provocation patterns, 41/53 (77.4%) had painDETECT scores ≤ 12, and 50/53 (94.3%) patients did not fulfill criteria for widespread pain. Thus, the included nsCLBP cohort predominantly presented with nociceptive pain features.

### 3.2. Indications for cohort-specific CPM differences

The median of the maximum pain perceived in response to the cold water bath did not differ between cohorts (p = 0.157, η^2^ = 0.02) and was 9 (7.0 - 9.0) for patients with nsCLBP, 9 (8.0 - 9.0) for patients with CRPS, 7 (4.5 - 9.0) for patients with painSCI, and 8 (6.3 - 9.0) for controls.

Low pain (< NRS 4/10) during the hand immersion into cold water was reported by 4 patients with nsCLBP, 3 patients with painSCI and 3 controls. These participants had no influence on the results as established from the influential case analysis. The PPTs for sequential CPM effects were assessed on average 3 min 06 s (standard deviation = 51 s) after the cold water bath.

CPM effects differed between cohorts (’timepoint X cohort’ interaction: F = 2.8, p = 0.011) (Figure 1, Table 2). Post-hoc tests indicated that patients with nsCLBP showed smaller parallel inhibitory CPM effects compared to patients with painSCI (t = -2.9, p = 0.043). Of note, statistical inference of the model changed after the removal of 13 influential cases (of 417 observations) and thus, the reported results refer to the model without influential cases. In the full model, the significant post-hoc test was only significant without multiple comparisons correction. The model also showed that males had higher PPTs compared to females (main effect of gender: F = 8.1, p = 0.005).

**Figure 1.**
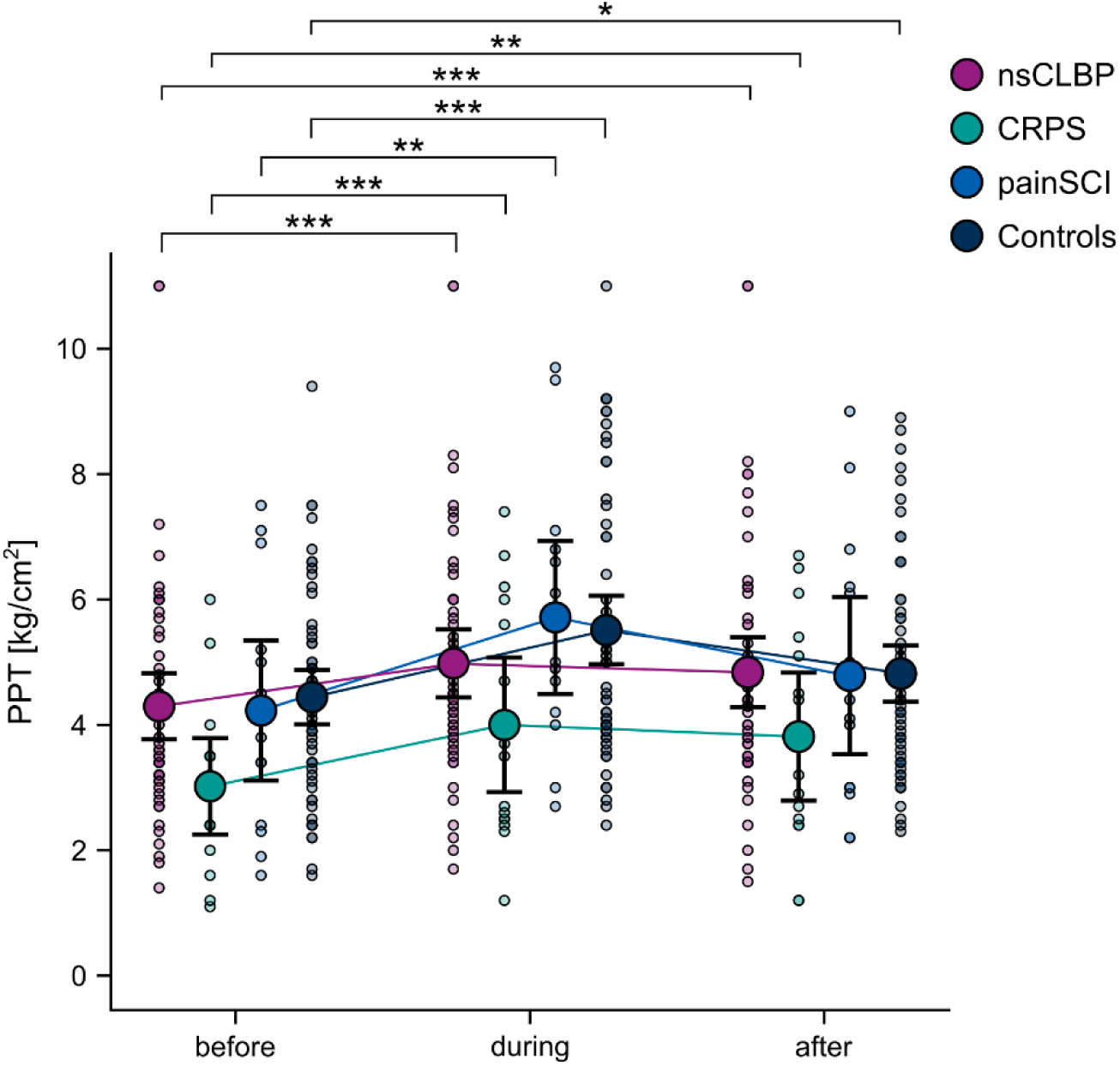
Cohort-specific CPM effects. Absolute PPT values at each timepoint of the CPM paradigm. The plots show the raw data (colored semi-transparent dots), means (colored dots) and 95% confidence intervals (black bars). Significances refer to post-hoc tests of linear mixed models performed for each cohort independently to examine whether each cohort showed significant CPM effects. \**p* < 0.05, \*\**p* < 0.01, \*\*\**p* < 0.001. CPM: conditioned pain modulation; CRPS: complex regional pain syndrome; nsCLBP: non-specific chronic low back pain; painSCI: neuropathic pain after spinal cord injury; PPT: pressure pain threshold.

**Table 2.**
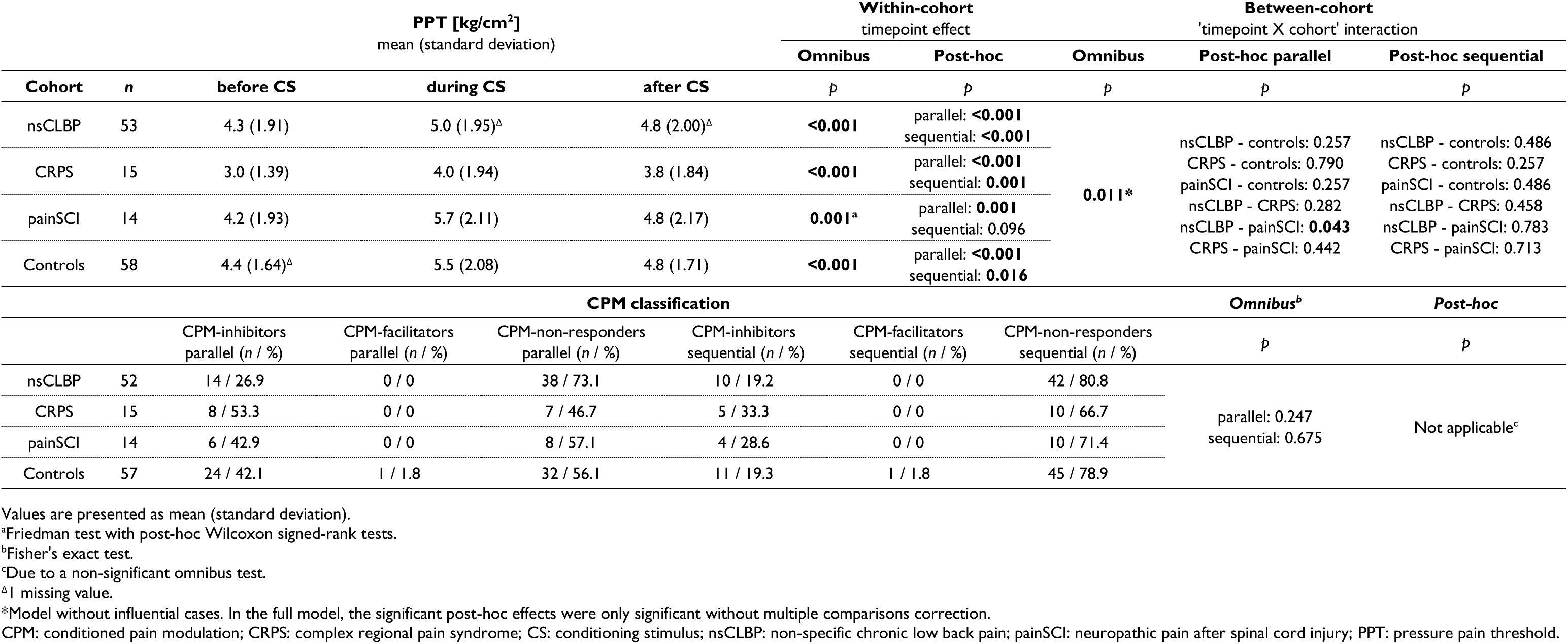
Cohort-specific CPM effects.

The model with PPT *changes* as the dependent variable and with correction for baseline PPTs showed qualitatively similar results (for details see Supplementary Results R1, Supplementary Figure S1, and Supplementary Table S1).

Baseline PPTs were significantly different between cohorts (χ^2^ = 8.9, p = 0.031, η^2^ = 0.04) with patients with CRPS showing lower baseline PPTs compared to patients with nsCLBP (p = 0.046, r = 0.30) and controls (p = 0.018, r = 0.35). Otherwise, no cohort differences in baseline PPTs were observed (p’s ≥ 0.221, r’s ≤ 0.30).

When examining CPM effects in each cohort separately, it was observed that all cohorts showed significant inhibitory CPM effects (F’s ≥ 11.0; Friedman test for patients with painSCI χ^2^ = 13.9; p’s ≤ 0.001). Parallel inhibitory CPM effects were observed in all cohorts (t’s ≥ 4.4; for patients with painSCI W = 3.0; p’s ≤ 0.001) and sequential inhibitory CPM effects in all cohorts (t’s ≥ 2.4, p’s ≤ 0.016) except for patients with painSCI (W = 25.5, p = 0.096) (Figure 1, Table 2). Male patients with nsCLBP showed higher PPTs compared to female patients with nsCLBP (F = 10.2, p = 0.002).

#### Associations of clinical pain characteristics and psychological factors with CPM effects

For the most part, there were no associations of CPM effects with clinical pain characteristics or psychological factors (Figure 2, Supplementary Table S2). Only when all patient cohorts were combined, a weak positive correlation between sequential inhibitory CPM effects and HADS depression scores was observed (rho = 0.27, p = 0.014). Of note, this correlation would not have persisted after multiple comparison correction. Rank-based regressions did not reveal associations that differed between cohorts (Supplementary Table S2).

**Figure 2.**
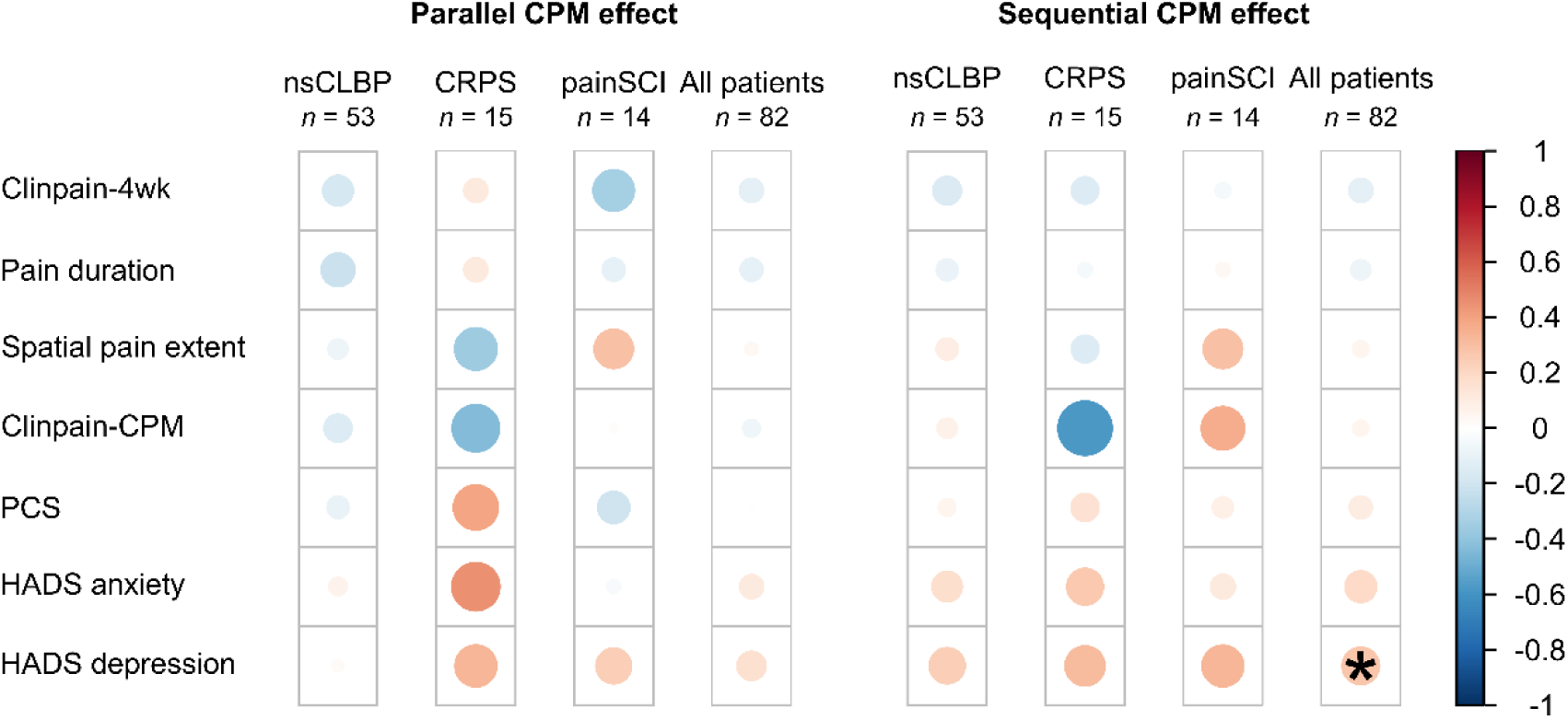
Associations of CPM effects with clinical pain characteristics and psychological factors. Correlation matrix displaying the strength of Spearman correlations between CPM effects (parallel and sequential) and clinical pain characteristics and psychological factors. Larger sizes and more intense colors of the circles indicate stronger correlations. Red shades indicate positive correlations and blue shades indicate negative correlations. Sample size for clinpain-4wk correlations: nsCLBP N = 52 due to 1 missing (implausible) value. Sample size for pain duration correlations: nsCLBP N = 52 due to 1 missing value (participant did not indicate month of pain onset). Sample size for clinpain-CPM correlations: nsCLBP N = 46, CRPS N = 12, painSCI N = 9 due to missing values. *p <0.05. Clinpain-4wk: average clinical pain intensity over the past 4 weeks; clinpain-CPM: within-CPM-session clinical pain; CPM: conditioned pain modulation; CRPS: complex regional pain syndrome; HADS: Hospital Anxiety and Depression Scale; nsCLBP: non-specific chronic low back pain; painSCI: neuropathic pain after spinal cord injury; PCS: Pain Catastrophizing Scale.

### 3.3. Subgroups with varying degrees of inhibitory CPM effects exist

The single-class latent class linear mixed model showed a significant effect of ‘timepoint’, (Wald = 106.6, p < 0.001) and a main effect of gender (Wald = 10.6, p = 0.001). Thus, timepoint and gender were both included as independent variables in the multi-class models.

Based on the BIC, the latent class linear mixed model with 3 classes, i.e., subgroups, was the best model (Table 3). All 3 subgroups showed high a confidence of subgroup membership (> 85.3%) and included more than 5% of the total sample (Table 3).

**Table 3.** Latent class linear mixed model fit and PPT-subgroup characteristics.

| PPT model | BIC | Posterior probabilities<br>Confidence of subgroup membership |  |  | Subgroup sizes |  |  |
| --- | --- | --- | --- | --- | --- | --- | --- |
|  |  | PPT-subgroup 1 | PPT-subgroup 2 | PPT-subgroup 3 | PPT-subgroup 1<br>(n / %) | PPT-subgroup 2<br>(n / %) | PPT-subgroup 3<br>(n / %) |
| 1 class | 1381.1 |  |  |  |  |  |  |
| 2 classes | 1348.1 |  |  |  |  |  |  |
| 3 classes | 1346.2 | 95.9 | 95.7 | 85.3 | 8 / 5.7 | 115 / 82.1 | 17 / 12.1 |
| 4 classes | 1353.9 |  |  |  |  |  |  |
|  |  | Cohort proportions in best model<br>(n / %) |  |  | Subgroup proportions in cohorts<br>[%] |  |  |
| nsCLBP |  | 2 / 25.0 | 43 / 37.4 | 8 / 47.1 | 3.8 | 81.1 | 15.1 |
| CRPS |  | 0 / 0 | 14 / 12.2 | 1 / 5.9 | 0 | 93.3 | 6.7 |
| painSCI |  | 2 / 25.0 | 11 / 9.6 | 1 / 5.9 | 14.3 | 78.6 | 7.1 |
| Controls |  | 4 / 50.0 | 47 / 40.9 | 7 / 41.2 | 6.9 | 81.0 | 12.1 |

BIC: Bayesian information criterion; CRPS: complex regional pain syndrome; nsCLBP: non-specific chronic low back pain; painSCI: neuropathic pain after spinal cord injury; PPT: pressure pain threshold.

With regards to CPM effects, the 3 subgroups are best described as follows (Figure 3, Table 4): PPT-subgroup 1 (N = 8), “low baseline PPT and large parallel inhibitory CPM effect”, PPT-subgroup 2 (N = 115), “low baseline PPT and small parallel and sequential inhibitory CPM effects”, and PPT-subgroup 3 (N = 17), “high baseline PPT and moderate parallel and sequential inhibitory CPM effects”. All subgroups included patients and controls in different proportions (Table 3).

**Figure 3.**
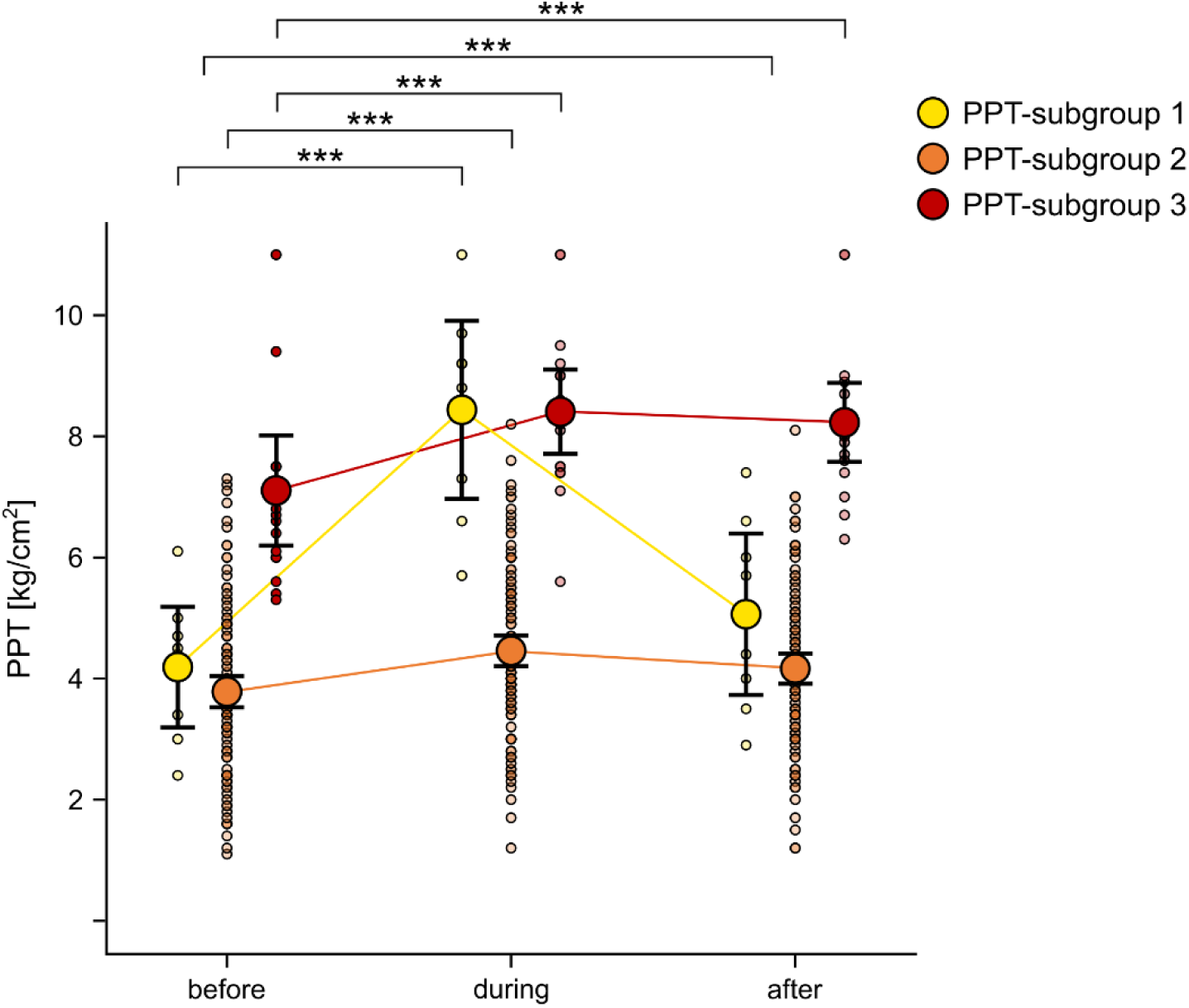
Subgroup-specific CPM effects. Absolute PPT values at each timepoint of the CPM paradigm for the 3 identified PPT-subgroups. The plots show the raw data (colored semi-transparent dots), means (colored dots) and 95% confidence intervals (black bars). Significances refer to post-hoc tests of linear mixed models performed for each PPT-subgroup independently to examine whether each PPT-subgroup showed significant CPM effects. \**p* < 0.05, \*\**p* < 0.01, \*\*\**p* < 0.001. CPM: conditioned pain modulation PPT: pressure pain threshold.

**Table 4.** PPT-subgroup-specific CPM effects.

| PPT [kg/cm<sup>2</sup>]<br>mean (standard deviation) |  |  |  |  | Within-subgroup<br>timepoint effect<sup>a</sup> |  | Between-subgroup<br>CPM effect comparison<sup>a</sup> |  |  |
| --- | --- | --- | --- | --- | --- | --- | --- | --- | --- |
|  |  |  |  |  | Omnibus | Post-hoc | Omnibus | Post-hoc parallel | Post-hoc sequential |
| PPT-subgroup | n | before CS | during CS | after CS | p | p | p | p | p |
| PPT-subgroup 1 | 8 | 4.2 (1.19) | 8.4 (1.76) | 5.1 (1.59) | 0.002<sup>b</sup> | parallel: 0.016<br>sequential: 0.058 | parallel:<br><0.001<br>sequential:<br>0.006 | Subg. 1 - Subg. 2: <0.001<br>Subg. 1 - Subg. 3: <0.001<br>Subg. 2 - Subg. 3: 0.010 | Subg. 1 - Subg. 2: 0.231<br>Subg. 1 - Subg. 3: 0.530<br>Subg. 2 - Subg. 3: 0.008 |
| PPT-subgroup 2 | 115 | 3.8 (1.38)<sup>Δ</sup> | 4.5 (1.37)<sup>Δ</sup> | 4.2 (1.34)<sup>Δ</sup> | <0.001 | parallel: <0.001<br>sequential: <0.001 |  |  |  |
| PPT-subgroup 3 | 17 | 7.1 (1.77) | 8.4 (1.36) | 8.2 (1.27) | <0.001 | parallel: <0.001<br>sequential: <0.001 |  |  |  |
| CPM classification |  |  |  |  |  |  |  | Omnibus<sup>c</sup> | Post-hoc parallel |
|  |  | CPM-inhibitors<br>parallel (n / %) | CPM-facilitators<br>parallel (n / %) | CPM-non-responders<br>parallel (n / %) | CPM-inhibitors<br>sequential (n / %) | CPM-facilitators<br>sequential (n / %) | CPM-non-responders<br>sequential (n / %) | p | p |
| PPT-subgroup 1 | 8 | 8 / 100.0 | 0 / 0 | 0 / 0 | 2 / 25.0 | 0 / 0 | 6 / 75.0 | parallel: 0.002<br>sequential: 0.878 | Subg. 1 - Subg. 2: 0.001<br>Subg. 1 - Subg. 3: 0.002<br>Subg. 2 - Subg. 3: 0.815 |
| PPT-subgroup 2 | 113 | 39 / 34.5 | 1 / 0.9 | 73 / 64.6 | 24 / 21.2 | 1 / 0.9 | 88 / 77.9 |  |  |
| PPT-subgroup 3 | 17 | 5 / 29.4 | 0 / 0 | 12 / 70.6 | 4 / 23.5 | 0 / 0 | 13 / 76.5 |  |  |

<sup>a</sup>Tested using separate linear models.
<sup>b</sup>Friedman test with post-hoc Wilcoxon signed-rank tests.
<sup>c</sup>Fisher's exact test.
<sup>Δ</sup>I missing value.
CPM: conditioned pain modulation; CS: conditioning stimulus; PPT: pressure pain threshold.

The model with PPT *changes* as dependent variable and with correction for baseline PPTs showed qualitatively similar results in that it detected 2 subgroups with different degrees of inhibitory CPM effects and with different proportions of patients and controls (for details see Supplementary Results R2, Supplementary Figure S2, and Supplementary Tables S3 and S4).

All PPT-subgroups showed significant inhibitory CPM effects (F’s ≥ 19.6; Friedman test for PPT-subgroup 1 χ^2^ = 13; p’s ≤ 0.002). Parallel inhibitory CPM effects were present in all PPT-subgroups (t’s ≥ 5.8; for PPT-subgroup 1 W = 0; p’s ≤ 0.016) and sequential inhibitory CPM effects in PPT-subgroups 2 and 3 (t’s ≥ 4.8, p’s < 0.001) but not PPT-subgroup 1, where only a trend toward a sequential inhibitory CPM effect was observed (W = 4, p = 0.058) (Figure 3, Table 4). In PPT-subgroup 2, males showed higher PPTs compared to females (F = 11.7, p < 0.001).

The PPT-subgroups differed in their parallel (F = 59.4, p < 0.001, partial η^2^ = 0.47) and sequential (F = 5.3, p = 0.006, partial η^2^ = 0.07) inhibitory CPM effects: PPT-Subgroup 1 showed larger parallel inhibitory CPM effects compared to PPT-subgroup 2 (t = 10.8, p < 0.001, d = 3.9) and to PPT-subgroup 3 (t = 7.6, p < 0.001, d = 3.3) and PPT-subgroup 2 showed smaller inhibitory CPM effects compared to PPT-subgroup 3 (t = -2.6, p = 0.010, d = 0.7). Thus, PPT-subgroup 2 showed the smallest parallel inhibitory CPM effects. Sequential inhibitory CPM effects were also smaller in PPT-subgroup 2, but only compared to PPT-subgroup 3 (t = -3.1, p = 0.008, d = 0.8). No difference in sequential inhibitory CPM effects was observed between PPT-subgroup 1 and PPT-subgroup 2 (t = 1.4, p = 0.231, d = 0.5) or PPT-subgroup 3 (t = -0.6, p = 0.530, d = 0.3).

### 3.4. CPM responder classification

The SEM for PPTs was 16.0% and thus, a PPT increase of 32.0% was considered a real change in CPM beyond the error of measurement.

Cohort-specific proportions of CPM-inhibitors, CPM-facilitators and CPM-non-responders for parallel and sequential CPM effects are reported in Table 2. There was no difference in proportions between cohorts (p’s ≥ 0.247).

The subgroup-specific proportions (for PPT-subgroups) are reported in Table 4. Proportions differed significantly between PPT-subgroups for parallel CPM effects (p = 0.002) with all members of PPT-subgroup 1 being classified as CPM-inhibitors, while PPT-subgroup 2 (p < 0.001 vs. PPT-subgroup 1) and PPT-subgroup 3 (p = 0.002 vs. PPT-subgroup 1) also comprised CPM-facilitators or CPM-non-responders. Proportions of CPM-inhibitors, CPM-facilitators and CPM-non-responders did not differ between PPT-subgroup 2 and PPT-subgroup 3 (p = 0.815). For sequential CPM effects, the PPT-subgroups did not differ in their proportions of CPM-inhibitors, CPM-facilitators and CPM-non-responders (p = 0.878).

Relative CPM effects expressed as percentage values are displayed in Figure 4A for cohorts and in Figure 4B for subgroups.

**Figure 4.**
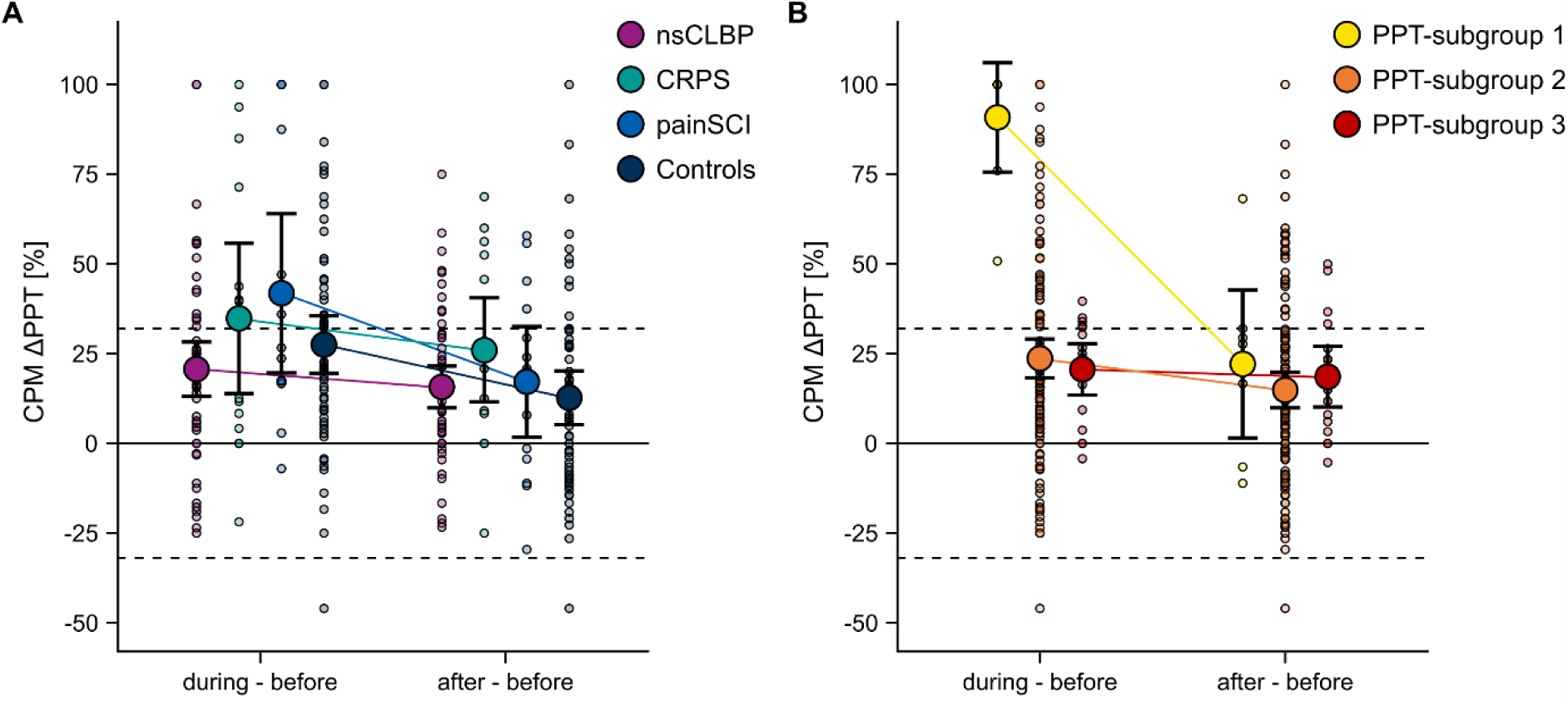
CPM responder classification for cohorts and subgroups. Relative PPT changes (CPM ΔPPT in percent) during the cold water bath (during - before, parallel CPM effect) and after the cold water bath (after - before, sequential CPM effect) for all cohorts (**A**) and for the 3 identified PPT-subgroups (**B**). Positive relative CPM ΔPPTs reflect inhibitory CPM effects (i.e., PPT increase) and negative values reflect facilitatory CPM effects (i.e., PPT decrease). The plots show the raw data (colored semi-transparent dots), means (colored dots) and 95% confidence intervals (black bars). The dotted lines reflect 2 SEM of the PPT (32.0%). CPM: conditioned pain modulation; CRPS: complex regional pain syndrome; nsCLBP: non-specific chronic low back pain; painSCI: neuropathic pain after spinal cord injury; PPT: pressure pain threshold.

### 3.5. Confounding factors

Proportions of participants in different menstrual cycle phases were not different between the cohorts (Supplementary Table S5) (p = 0.638) and thus, data were pooled to assess whether the menstrual cycle phase had an influence on CPM effects. There was no influence of menstrual cycle phase on CPM effects (Supplementary Table S5).

Proportions of participants with and without pain-relevant regular medication intake were different between the cohorts (p < 0.001). All patient cohorts reported regular pain-relevant medication intake more frequently than controls (all p’s ≤ 0.011) and patients with CRPS more frequently than patients with nsCLBP (p = 0.003) (Supplementary Table S6). Thus, cohorts were examined separately to assess whether the regular medication intake had an influence on CPM effects. Neither cohort showed an influence of regular medication intake on CPM effects (Supplementary Table S6).

## 4. Discussion

The present study aimed to compare CPM effects across patients with distinct chronic pain disorders, namely nsCLBP, CRPS, and painSCI, and pain-free controls, assessed using the same CPM paradigm. Cohort-specific differences in CPM effects were observed, mainly driven by smaller parallel inhibitory CPM effects in patients with nsCLBP compared to patients with painSCI. CPM effects were, to the largest part, not associated with the patients’ clinical pain characteristics or psychological features. Additionally, independent of cohort membership, the present study identified 3 subgroups with varying degrees of inhibitory CPM effects.

Deficient CPM is considered a common feature of various chronic pain disorders [22,44,74]. The present results do not support this notion, as patients with nsCLBP, CRPS, and painSCI did not show different CPM effects than controls. Only in patients with nsCLBP, reduced inhibitory CPM effects compared to patients with painSCI were observed. While studies reporting deficient CPM in nsCLBP cohorts compared to controls exist [7,50,62], others did not describe such differences [15,20,38,40,49,54,57]. A recent meta-analysis [47] highlighted that deficient CPM might depend on pain characteristics: deficient CPM was only detected in patients with CLBP reporting clinical pain intensities > NRS 5. In the present study, clinical pain intensities of the nsCLBP cohort were lower (median: NRS 4). Additionally, 94.3% of the included nsCLBP cohort had localized pain which has been associated with more effective CPM compared to patients with widespread CLBP [18]. Overall, the included nsCLBP cohort exhibited predominantly nociceptive pain features and according to current expert consensus, deficient CPM is not considered a characteristic associated with nociceptive pain mechanisms [71]. These aspects might explain the absence of a pronounced deficient CPM in the present nsCLBP cohort. Nevertheless, inhibitory CPM effects were smaller in patients with nsCLBP compared to painSCI, suggesting that deficient descending pain inhibition might play a more important role in musculoskeletal compared to neuropathic pain disorders.

Patients with CRPS and painSCI did not present with reduced inhibitory CPM effects. For CRPS, this is in line with existing literature [39]. Notably, the CRPS cohort included in the present study showed functional CPM despite displaying lower PPTs before the CS compared to controls. This indicates generalized pain hypersensitivity independent of impaired descending pain inhibition. In patients with painSCI, two studies reported deficient above-level CPM compared to controls [2,26]. This inconsistency with the present results might again be attributed to higher clinical pain intensities (mean: NRS 6.3 [2] and 5.5 [26]) than observed in the present study (median: NRS 4). In line with this notion, higher spontaneous pain intensity has been shown to correlate with smaller inhibitory CPM effects in painSCI [2,46]. An alternative explanation is the use of a thermal, rather than a mechanical TS. Thermal stimuli are susceptible to adaptation or habituation [24] which might contribute to TS increases over time unrelated to the painfulness of the CS. Another painSCI study [25] reported deficient CPM in the patients, but only for *at-level* CPM and not for *above-level* CPM effects, in line with the present results.

In an attempt to reduce the influences of baseline PPTs on the linear mixed model analyses, cohort-specific CPM differences were also investigated for PPT changes while correcting for baseline PPTs. Yet, these analyses did not offer additional insights. This is not surprising given that CPM effects, calculated as B - A, will, from a mathematical perspective, always depend on A, i.e., the baseline PPT. Importantly, this holds particularly true for relative CPM effects. For instance, the same absolute change will result in a larger relative change for lower baseline values. This effect is evident in the present CRPS cohort which showed qualitatively similar absolute CPM effects compared to controls (Figure 1B, Table 2), but larger relative CPM effects (Figure 4A) due to significantly lower baseline PPTs. Participants with higher baseline values might additionally experience ceiling effects due to safety cut-offs for applied TS intensities (e.g., 10kg for PPT in the present study). Therefore, CPM effects should not be interpreted without consideration of baseline TS values and caution is advised when comparing absolute with relative CPM effects. This is particularly important when CPM effects are assessed in painful, already sensitized areas of the patients [12] and favors CPM assessments in pain-free areas in which baseline pain sensitivity might be more comparable between cohorts.

Clinical pain characteristics did not correlate with CPM effects. This contradicts some disorder-specific observations, for example deficient CPM effect being associated with higher pain intensity in CLBP [47], longer pain duration in CLBP [47] and painSCI [17], and with more widespread pain in CLBP [18], painSCI [26], and fibromyalgia [32]. However, across chronic pain disorders, most studies did not report such relationships (reviewed in [12]). This was particularly true for studies using mechanical TS’s, which aligns with the present results. The associations might further depend on the type of pain investigated. In our previous study on the same painSCI cohort [46] and in the study by Albu and colleagues [2], CPM effects correlated with spontaneous pain intensity, while the present study used average pain intensities over the past 4 weeks for comparability across all included disorders.

For associations of psychological factors with CPM effects, larger sequential CPM effects correlated with higher HADS depression scores in the combined patient sample. However, given the correlation’s small effect size and the large number of performed correlational analyses, this result might reflect a spurious finding.

CPM effects were also independent of age, gender, and menstrual cycle phases, supporting previous studies in which CPM variability could not be explained by these factors [21,69,79,80]. Furthermore, a potential confounding by regular pain-relevant medication intake [19] was excluded.

The high inter-individual variability of CPM effects in pain-free controls [21,59] and chronic pain disorders [41,59] raises the question whether deficient CPM might not be a cohort- but rather a subgroup-specific or individual feature. In the present study, three homogenous subgroups showing different CPM effects were detected across all chronic pain disorders and controls. The subgroups showed significant inhibitory CPM effects and contained members of all cohorts, except for the most inhibitory subgroup which lacked patients with CRPS. No subgroup without inhibitory CPM effects was identified. Previous studies using hierarchical clustering have reported chronic pain patient subgroups with deficient CPM [36,55]. However, both studies performed the clustering exclusively on the patients and compared the resulting subgroups to controls. This approach neglects that also within pain-free cohorts, a spectrum from inhibitory to facilitatory CPM effects exists [30,59,69] and might artificially create differences between patient subgroups and controls.

Deficient CPM has been suggested as a predictor for chronic pain development [10,83] and pharmacological treatment efficacy [11,85]. Yet, if *groups* with deficient CPM remain undetected in a large, heterogenous sample, reliably identifying *individuals* with deficient CPM is doubtful. This is underlined by the observation that only 1 individual (of 140) was identified as a CPM-facilitator when accounting for the SEM of the TS [8,34]. Combining CPM with other, complementary, pain modulatory measures, e.g., temporal summation of pain as a pronociceptive measure [23,84], might be necessary to identify individuals with impaired endogenous pain modulation.

One limitation of the present study is that data of the CPM_SHAM_ paradigm could not be implemented in the linear mixed model analyses due to sample size limitations. However, choosing a reliable TS-CS combination, e.g., PPT and a cold water bath, might be clinically more relevant given that time constraints render the standard implementation of CPM_SHAM_ paradigms difficult. Additionally, the larger nsCLBP sample size might have favored the detection of deficient CPM in this cohort. However, the CRPS and painSCI samples are comparable to previous studies [2,26,39] and neither cohort showed a trend toward deficient CPM. Additionally, the sample analyzed in the present study included mostly white, European participants despite study participation being open to all ethnicities. This limits the present study’s generalizability and future work should aim to foster inclusion of participants of other ethnicities, for example by reaching out to respective advocacy groups.

In conclusion, comparing CPM effects measured by the same CPM paradigm among distinct chronic pain disorders, namely nsCLBP, CRPS, and neuropathic pain after SCI, and pain-free controls demonstrated reduced inhibitory CPM effects exclusively in patients with nsCLBP. CPM variability remained largely unexplained by clinical pain characteristics or psychological factors. Across the three chronic pain disorders and controls, 3 subgroups including patients and controls with varying degrees of inhibitory CPM effects were identified. These findings oppose the notion of deficient descending pain inhibition as common feature of chronic pain disorders. Moreover, they reinforce the importance of further investigating sources of CPM variability and multimodal assessments of pain modulation, and they challenge whether individual CPM effects are useful for predicting individuals at risk of progressing to chronic pain or informing treatment decisions.

## Data Availability

All data produced in the present study are available upon reasonable request to the authors.

## Data statement

Code and data used in this manuscript will be made publicly available. However, only a reduced dataset can be shared because not all participants have agreed on further use of their data.

## Author contributions

Conceptualization: L.S., I.D.S., R.L., J.R., M.B., B.W., M.H., and P.S.; Data curation: L.S., I.D.S., L.G., and R.L.; Formal analysis: L.S.; Funding acquisition: F.B., M.H. and P.S.; Investigation: L.S., I.D.S., L.G., R.L., A.L., F.B., J.R., M.B., B.W., and P.S.; Methodology: L.S., I.D.S., R.L., F.B., J.R., M.B., B.W., M.H., and P.S.; Project administration: L.S., I.D.S., M.H., and P.S.; Supervision; M.H. and P.S.; Visualization: L.S.; Writing - original draft; L.S.; and Writing - review and editing: L.S., I.D.S, L.G., R.L., A.L., J.R., B.W., M.H., and P.S..

## Acknowledgments

We thank all participants who took part in the study. Additionally, we thank Lucas Tauschek and Simon Carisch for their support during data acquisition and László Demkó for setting up the custom-made software for spatial pain extent calculation. We further thank Balgrist Campus for their support and providing the infrastructure to perform the present study.

