## Supplementary Material for "Challenging deficient inhibitory conditioned pain modulation as common chronic pain feature and detectable subgroup characteristic"

#### Supplementary Methods

##### M1. PPT *change* models

For the PPT *change* analysis, the linear mixed model was performed with the PPT *change* (i.e., during - before and after - before) as the dependent variable and the independent variables timepoint (2 levels: 'during-before', 'after-before'), cohort (4 levels: 'nsCLBP', 'CRPS', 'painSCI', 'controls'), baseline PPT, and the interaction of interest, i.e., 'timepoint X cohort'. Post-hoc tests were performed using planned comparisons (all 'timepoint X cohort' interactions except for 'timepoint during' vs. 'timepoint after' comparisons).

### Supplementary Results

#### R1. PPT *change* model: Cohort-specific approach

The model with PPT *changes* as the dependent variable and with correction for baseline PPTs showed that the relationship between parallel and sequential CPM effects varied between cohorts ('timepoint X cohort' interaction: $F = 5.3$, $p = 0.002$) (Supplementary Figure S1, Supplementary Table S1). Post-hoc tests showed that patients with nsCLBP presented with the smallest decrease from parallel to sequential CPM effects which was significantly less pronounced than both the decrease observed in patients with painSCI ($t = 2.8$, $p = 0.016$) and in controls ($t = 3.4$, $p = 0.005$). Of note, statistical inference of the model changed after removal of 7 influential cases (of 276 observations) and thus, the reported results refer to the model without influential cases. In the full model, the significant post-hoc tests were only significant without multiple comparisons correction.

#### R2. PPT *change* model: Subgroup-specific approach

For the model with PPT *changes* as the dependent variable and with correction for baseline PPTs, the single-class latent class linear mixed model showed a significant effect of 'timepoint' (Wald = 23.0, $p < 0.001$). Thus, timepoint and baseline PPTs were included as independent variables in the multi-class models.

Based on the BIC, the latent class linear mixed model with 2 classes, i.e., subgroups, was the best model (Supplementary Table S3). Both subgroups showed a high confidence of subgroup membership (> 96.7%) and included more than 5% of the total sample (Supplementary Table S3).

With regards to CPM effects, the 2 subgroups are best described as follows (Supplementary Figure S2, Supplementary Table S4): PPT *change*-subgroup 1 (N = 130), "small parallel and small sequential CPM effects", and PPT *change*-subgroup 2 (N = 8) "large parallel and small sequential CPM effects". Proportions of cohorts in the respective subgroups are described in Supplementary Table S3.

### Supplementary Figures

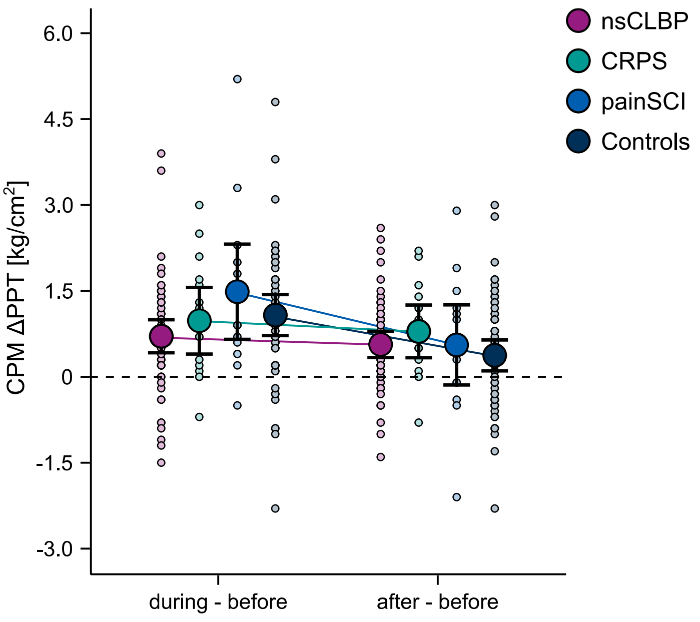


**Figure S1. Cohort-specific CPM effects: differences in PPT *changes*.** Absolute PPT changes (CPM ΔPPT) during the cold water bath (during - before, parallel CPM effect) and after the cold water bath (after - before, sequential CPM effect). Positive absolute CPM ΔPPTs reflect inhibitory CPM effects (i.e., PPT increase) and negative values reflect facilitatory CPM effects (i.e., PPT decrease). The plots show the raw data (light-colored dots), means (colored dots) and 95% confidence intervals (black bars). CPM: conditioned pain modulation; CRPS: complex regional pain syndrome; nsCLBP: non-specific chronic low back pain; painSCI: neuropathic pain after spinal cord injury; PPT: pressure pain threshold.

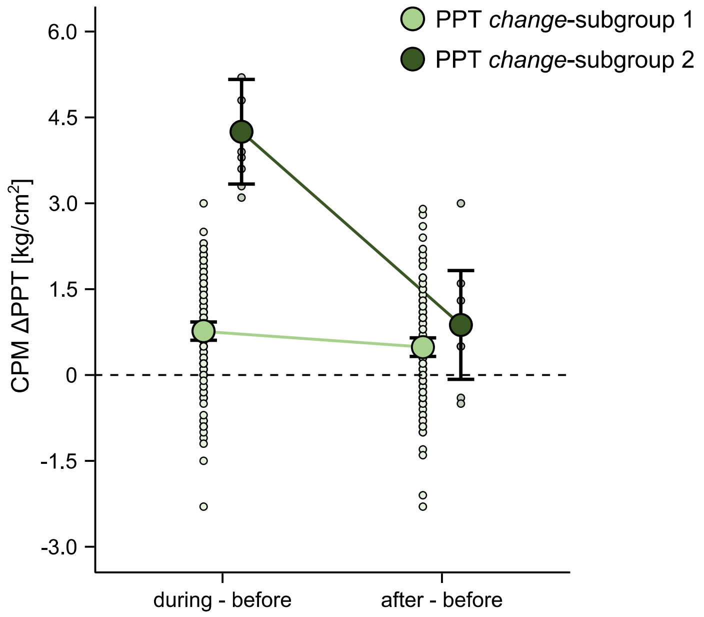


**Figure S2. Subgroup-specific CPM effects: differences in PPT *changes*.** Absolute PPT changes (CPM ΔPPT) during the cold water bath (during - before, parallel CPM effect) and after the cold water bath (after - before, sequential CPM effect) for the 2 identified PPT *change*-subgroups. Positive absolute CPM ΔPPTs reflect inhibitory CPM effects (i.e., PPT increase) and negative values reflect facilitatory CPM effects (i.e., PPT decrease). The plots show the raw data (light-colored dots), means (colored dots) and 95% confidence intervals (black bars). CPM: conditioned pain modulation PPT: pressure pain threshold.

### Supplementary Tables

**Table S1 Cohort-specific CPM effects: differences in PPT *changes*.**

| | | PPT change [kg/cm$^2$] mean (standard deviation) | | Between-cohort 'timepoint X cohort' interaction | |
|---|---|---|---|---|---|
| | | | | **Omnibus** | **Post-hoc** |
| | **n** | during - before | after - before | *p* | *p* |
| nsCLBP | 52 | 0.7 (1.04) | 0.6 (0.82) | **0.002*** | nsCLBP - controls: **0.005**<br>CRPS - controls: 0.160<br>painSCI - controls: 0.506<br>nsCLBP - CRPS: 0.506<br>nsCLBP - painSCI: **0.016**<br>CRPS - painSCI: 0.134 |
| CRPS | 15 | 1.0 (1.05) | 0.8 (0.83) | | |
| painSCI | 14 | 1.5 (1.44) | 0.6 (1.21) | | |
| Controls | 57 | 1.1 (1.35) | 0.4 (1.02) | | |

Values are presented as mean (standard deviation).
*Model without influential cases. In the full model, the significant post-hoc effects were only significant without multiple comparisons correction.
CPM: conditioned pain modulation; CRPS: complex regional pain syndrome; CS: conditioning stimulus; nsCLBP: non-specific chronic low back pain; painSCI: neuropathic pain after spinal cord injury; PPT: pressure pain threshold.

**Table S2 Results from rank-based regressions used to test cohort differences in associations of CPM effects with clinical pain characteristics and psychological factors.**

| | Spearman correlations *rho / p* | | | | Rank-based regression cohort interaction effect | |
|---|---|---|---|---|---|---|
| | **nsCLBP (*n* = 53)** | **CRPS (*n* = 15)** | **painSCI (*n* = 14)** | **All patients (*n* = 82)** | ***F*** | ***p*** |
| **Parallel CPM effects** | | | | | | |
| Clinpain-4wk[a] | -0.18 / 0.214 | 0.11 / 0.690 | -0.33 / 0.243 | -0.11 / 0.316 | 0.6 | 0.577 |
| Pain duration[b] | -0.22 / 0.127 | 0.11 / 0.689 | -0.10 / 0.731 | -0.10 / 0.370 | 0.1 | 0.924 |
| Spatial pain extent | -0.08 / 0.597 | -0.35 / 0.198 | 0.30 / 0.299 | 0.03 / 0.768 | 1.3 | 0.271 |
| Clinpain-CPM[c] | -0.15 / 0.331 | -0.44 / 0.157 | 0.02 / 0.965 | -0.06 / 0.630 | 0.8 | 0.458 |
| PCS | -0.09 / 0.516 | 0.39 / 0.149 | -0.20 / 0.492 | -0.01 / 0.958 | 1.2 | 0.310 |
| HADS anxiety | 0.06 / 0.656 | 0.45 / 0.090 | -0.04 / 0.900 | 0.11 / 0.318 | 0.8 | 0.466 |
| HADS depression | 0.03 / 0.855 | 0.34 / 0.216 | 0.25 / 0.394 | 0.16 / 0.144 | 0.4 | 0.701 |
| **Sequential CPM effects** | | | | | | |
| Clinpain-4wk[a] | -0.15 / 0.281 | -0.15 / 0.606 | -0.05 / 0.871 | -0.11 / 0.321 | 0.5 | 0.634 |
| Pain duration[b] | -0.09 / 0.538 | -0.04 / 0.882 | 0.04 / 0.893 | -0.08 / 0.495 | 0.7 | 0.520 |
| Spatial pain extent | 0.09 / 0.516 | -0.15 / 0.599 | 0.30 / 0.299 | 0.05 / 0.646 | 1.4 | 0.252 |
| Clinpain-CPM[c] | 0.08 / 0.617 | -0.57 / 0.052 | 0.36 / 0.336 | 0.05 / 0.689 | 2.6 | 0.086 |
| PCS | 0.06 / 0.694 | 0.15 / 0.589 | 0.08 / 0.773 | 0.10 / 0.370 | 0 | 0.970 |
| HADS anxiety | 0.18 / 0.215 | 0.26 / 0.349 | 0.12 / 0.686 | 0.19 / 0.082 | 0.1 | 0.908 |
| HADS depression | 0.25 / 0.078 | 0.31 / 0.259 | 0.34 / 0.239 | **0.27 / 0.014** | 0.2 | 0.818 |

[a]NsCLBP *N* = 52 due to 1 missing (implausible) value.
[b]NsCLBP *N* = 52 due to 1 missing value (participant did not indicate month of pain onset).
[c]NsCLBP *N* = 46, CRPS *N* = 12, painSCI *N* = 9 due to missing values.
Clinpain-4wk: average clinical pain intensity over the past 4 weeks; clinpain-CPM: within-CPM-session clinical pain; CPM: conditioned pain modulation; CRPS: complex regional pain syndrome; HADS: Hospital Anxiety and Depression Scale; nsCLBP: non-specific chronic low back pain; painSCI: neuropathic pain after spinal cord injury; PCS: Pain Catastrophizing Scale;

**Table S3 Latent class linear mixed model fit and PPT *change*-subgroup characteristics.**

| | | **Posterior probabilities** Confidence of subgroup membership | | **Subgroup sizes** | |
|---|---|---|---|---|---|
| **PPT *change* model** | **BIC** | **PPT *change*-subgroup 1** | **PPT *change*-subgroup 2** | **PPT *change*-subgroup 1** (*n* / %) | **PPT *change*-subgroup 2** (*n* / %) |
| 1 class | 820.0 | | | | |
| 2 classes | 782.9 | 99.3 | 96.7 | 130 / 94.2 | 8 / 5.8 |
| 3 classes | 794.2 | | | | |
| | | **Cohort proportions in best model** (n / %) | | **Subgroup proportions in cohorts** [%] | |
| nsCLBP | | 50 / 38.5 | 2 / 25.0 | 94.3 | 3.8 |
| CRPS | | 15 / 11.5 | 0 / 0 | 100.0 | 0 |
| painSCI | | 12 / 9.2 | 2 / 25.0 | 85.7 | 14.3 |
| Controls | | 53 / 40.8 | 4 / 50.0 | 91.4 | 6.9 |

BIC: Bayesian information criterion; CRPS: complex regional pain syndrome; nsCLBP: non-specific chronic low back pain; painSCI: neuropathic pain after spinal cord injury; PPT: pressure pain threshold.

**Table S4 PPT *change*-subgroup-specific CPM effects.**

| | | **PPT change [kg/cm$^2$]** mean (standard deviation) | |
|---|---|---|---|
| **PPT *change*-subgroup** | ***n*** | during - before | after - before |
| PPT *change*-subgroup 1 | 130 | 0.8 (0.91) | 0.5 (0.94) |
| PPT *change*-subgroup 2 | 8 | 4.3 (1.09) | 0.9 (1.14) |

CPM: conditioned pain modulation; PPT: pressure pain threshold.

**Table S5 Menstrual cycle phase proportions across cohorts and CPM effects in different menstrual cycle phases.**

| | | Menstrual cycle phase | | | | Across-cohort proportions | |
|---|---|---|---|---|---|---|---|
| | | | | | | Omnibus[a] | Post-hoc |
| **Cohort** | ***n*** | menstruation (*n* / %) | follicular (*n* / %) | ovulation (*n* / %) | luteal (*n* / %) | *p* | *p* |
| nsCLBP | 14 | 3 / 21.4 | 5 / 35.7 | 2 / 14.3 | 4 / 28.6 | 0.638 | Not applicable[c] |
| CRPS | 6 | 3 / 50.0 | 0 / 0 | 0 / 0 | 3 / 50.0 | | |
| painSCI | 0 | | | | | | |
| Controls | 13 | 3 / 23.1 | 3 / 23.1 | 2 / 15.4 | 5 / 38.5 | | |

| | | PPT change [kg/cm²] median (interquartile range) | | Between-menstrual cycle phases timepoint effect | |
|---|---|---|---|---|---|
| | | | | **Omnibus[b]** | **Post-hoc** |
| | | during - before | after - before | *p* | *p* |
| Menstruation | 9 | 0.9 (0.30 - 1.60) | 0.6 (0.30 - 0.90) | parallel: 0.864<br>sequential: 0.603 | Not applicable[c] |
| Follicular | 8 | 1.0 (0.28 - 1.33) | 0.2 (-0.03 - 0.35) | | |
| Ovulation | 4 | 0.7 (0.08 - 0.90) | 0.5 (-0.58 - 1.33) | | |
| Luteal | 12 | 1.3 (0.18 - 1.80) | 0.5 (0.25 - 0.85) | | |

[a]Fisher's exact test.
[b]Kruskal-Wallis test due to small menstrual cycle phase subgroups.
[c]Due to a non-significant omnibus test.
CPM: conditioned pain modulation; CRPS: complex regional pain syndrome; nsCLBP: non-specific chronic low back pain; painSCI: neuropathic pain after spinal cord injury; PPT: pressure pain threshold.

**Table S6 Regular medication intake proportions across cohorts and CPM effects in participants with and without regular medication intake.**

| | | Regular medication intake | | | | | | | Across-cohort proportions | |
|---|---|---|---|---|---|---|---|---|---|---|
| | | | | | | | | | **Omnibus**[a] | **Post-hoc** |
| **Cohort** | ***n*** | M01A (*n* / %) | N02 (*n* / %) | N03 (*n* / %) | N05 (*n* / %) | N06 (*n* / %) | yes (*n* / %) | no (*n* / %) | *p* | *p* |
| nsCLBP | 53 | 6 / 11.3 | 1 / 1.9 | 2 / 3.8 | 4 / 7.5 | 7 / 13.2 | 12 / 22.6 | 41 / 77.4 | **<0.001** | nsCLBP - controls: **0.011**<br>CRPS - controls: **<0.001**<br>painSCI - controls: **0.006**<br>nsCLBP - CRPS: **0.003**<br>nsCLBP - painSCI: 0.322<br>CRPS - painSCI: 0.143 |
| CRPS | 15 | 2 / 13.3 | 8 / 53.3 | 4 / 26.7 | 0 / 0 | 6 / 40.0 | 10 / 66.7 | 5 / 33.3 | | |
| painSCI | 14 | 0 / 0 | 1 / 7.1 | 5 / 35.7 | 0 / 0 | 1 / 7.1 | 5 / 35.7 | 9 / 64.3 | | |
| Controls | 58 | 0 / 0 | 1 / 1.7 | 0 / 0 | 1 / 1.7 | 1 / 1.7 | 3 / 5.2 | 55 / 94.8 | | |

| **Regular medication intake** | ***n*** | **PPT change [kg/cm²]** median (interquartile range) | | **With or without regular medication intake** timepoint effect | |
|---|---|---|---|---|---|
| | | | | **Omnibus**[b] | |
| | | during - before | after - before | *p* | Effect size |
| nsCLBP - yes | 12 | 0.5 (-0.53 - 1.35) | 0.6 (0.18 - 1.13) | parallel: 0.487<br>sequential: 0.625 | parallel: r = 0.10<br>sequential: r = 0.07 |
| nsCLBP - no | 41 | 0.8 (0.28 - 1.23)[Δ] | 0.5 (0.08 - 1.00)[Δ] | | |
| CRPS - yes | 10 | 1.3 (0.40 - 1.68) | 1.0 (0.45 - 1.50) | parallel: 0.198<br>sequential: 0.177 | parallel: r = 0.35<br>sequential: r = 0.36 |
| CRPS - no | 5 | 0.2 (0 - 0.70) | 0.5 (0 - 0.50) | | |
| painSCI - yes | 5 | 0.9 (0.60 - 1.80) | 0.3 (-0.40 - 1.10) | parallel: 0.640<br>sequential: 0.423 | parallel: r = 0.14<br>sequential: r = 0.23 |
| painSCI - no | 9 | 1.4 (0.70 - 2.00) | 0.6 (-0.10 - 1.50) | | |
| Controls - yes | 3 | 2.1 (0.85 - 2.20) | -0.1 (-0.30 - 0.20) | parallel: 0.474<br>sequential: 0.442 | parallel: r = 0.10<br>sequential: r = 0.10 |
| Controls - no | 55 | 0.9 (0.50 - 1.58)[Δ] | 0.4 (-0.40 - 1.10)[Δ] | | |

[a]Fisher's exact test.
[b]Wilcoxon rank-sum test
[c]Due to a non-significant omnibus test.
[Δ]1 missing value.
CPM: conditioned pain modulation; CRPS: complex regional pain syndrome; nsCLBP: non-specific chronic low back pain; painSCI: neuropathic pain after spinal cord injury; PPT: pressure pain threshold.
